# The Unsteady Return of Command-Following: Recovery and Instability of Bedside Motor Command-Following After Acute Brain Injury

**DOI:** 10.64898/2026.06.19.26356104

**Authors:** Alon Gorenshtein, Yosef Adiniaev, Mahmud Omar, Yiftach Barash, Eyal Klang, Oved Daniel

**Affiliations:** Department of Neurology, Beth Israel Deaconess Medical Center, Harvard Medical School, Boston, MA, USA; BRIDGE GenAI Lab, Beth Israel Deaconess Medical Center, Boston, MA, USA; The Windreich Department of Artificial Intelligence and Human Health, Mount Sinai Medical Center, NY, USA; Department of Radiology, Beth Israel Deaconess Medical Center, Harvard Medical School, Boston, MA, USA; Neurology Division, Tel Aviv Sourasky University Medical Center, Tel Aviv, Israel

**Keywords:** disorders of consciousness, command-following, Glasgow Coma Scale, acute brain injury, competing risks, sedation, neurocritical care

## Abstract

**Background/Objective:** Following a verbal command marks the bedside transition from unresponsiveness to overt recovery of consciousness after acute brain injury. Its timing across phenotypes, stability once present, and dependence on sedation are uncharacterized at scale.

**Methods:** Retrospective cohort of adults with acute brain injury, first intensive care unit stay, MIMIC-IV. Command-following was the Glasgow Coma Scale motor response ”Obeys Commands.” Among patients not following commands at admission, cumulative incidence was estimated with death or hospice and discharge without recovery as competing events. Instability was quantified as transient first recovery and threshold crossings; examinations were tagged for concurrent sedation. Principal findings were externally validated in the multicenter eICU Collaborative Research Database.

**Results:** Of 13,900 brain-injured patients with three or more motor examinations, 5,498 (39.6%) were not following commands at admission. The cumulative incidence of first command-following was 43.5% by 24 hours and 65.0% by 14 days, ranging at 14 days from 36.9% in anoxic injury to 77.2% in ischemic stroke (anoxic versus ischemic stroke at 72 hours, difference 0.41; adjusted P = .002). Among 3,573 patients who recovered, the first recovery was transient in 22.2%, and 62.4% crossed the threshold repeatedly. Non-following was strongly associated with sedation, consistent with an arousal-dependent examination. In eICU, the 14-day incidence was 64.8%, and transient first recovery was 22.7%, closely matching the primary cohort.

**Conclusions:** After acute brain injury, overt bedside command-following returns early but unsteadily, with phenotype-dependent timing, threshold fluctuation, and strong dependence on sedation. A single charted observation is an unreliable index of the underlying state.

## Introduction

The ability to follow a spoken command separates the unresponsive states (coma and the vegetative or unresponsive wakefulness state) from the recovery of consciousness, and within the minimally conscious state it marks the transition from reflexive to purposeful behavior.[1,2,3] In the intensive care unit (ICU), the motor component of the Glasgow Coma Scale records this ability many times each day, and its return is both the first available evidence that consciousness is recovering and the signal clinicians use to time conversations about prognosis and goals of care.[4,5]

Two features of this examination complicate its interpretation, and both are well described in the rehabilitation setting but rarely quantified during the acute illness. First, responsiveness in the minimally conscious state fluctuates, so that the same patient follows commands at one assessment and not at the next; this instability is a principal reason that bedside diagnosis of consciousness is wrong in roughly 4 of 10 specialist examinations.[6,7] Second, the examination is suppressed by the sedation and analgesia administered to most mechanically ventilated brain-injured patients, so that an absent response can reflect the drug rather than the injury.[8,9] Whether command-following, when it first returns after acute brain injury, is stable or fluctuating, and how much of its timing is attributable to sedation, has not been described at the scale of routine care.

Prior work has measured the time to follow commands in traumatic brain injury and shown that a longer interval predicts worse functional recovery, but this evidence is confined to a single phenotype, treats command-following as a predictor rather than as an outcome to be characterized, and does not address the competing risk of death or withdrawal of care that removes patients from observation before recovery can occur.[10,11] We used a large critical care database to describe, across six acute brain-injury phenotypes and with a competing-risks design, when overt bedside command-following first returns, how stable it is once it appears, and how it relates to sedation depth, and we tested how much of the eventual recovery is already legible in the first day of bedside behavior. Throughout, the endpoint is the overt, charted motor response, which is what clinicians act on; covert consciousness and cognitive-motor dissociation are not detectable from this signal and are not the object of study.

## Methods

### Study Design and Setting

This was a retrospective cohort study of MIMIC-IV version 3.1, a single-center critical care database of adults admitted to the ICUs of Beth Israel Deaconess Medical Center between 2008 and 2019.[12] Reporting followed STROBE and RECORD, and the prediction component followed TRIPOD.[13,14,15] All analyses were descriptive or methodological; no causal or treatment-effect claim was made.

### Cohort Identification

Adults (age >= 18 years) with acute brain injury were eligible, defined from International Classification of Diseases, Ninth and Tenth Revision diagnosis codes in any position (not only the principal diagnosis) and grouped into six phenotypes: acute ischemic stroke, traumatic brain injury, intracerebral hemorrhage, subarachnoid hemorrhage, subdural hemorrhage, and anoxic injury (code lists, eTable 1). Any-position coding maximized sensitivity, because the brain injury is frequently not the principal code (anoxic injury, for example, is usually secondary to a cardiac-arrest principal diagnosis); the specificity trade-off is addressed in the Limitations. When phenotypes co-occurred (11.4% of hospitalizations), a single primary phenotype was assigned by a fixed clinical priority hierarchy favoring the more procedurally specific and acute diagnosis (subarachnoid, then intracerebral, subdural, traumatic, ischemic, anoxic), with a single-phenotype sensitivity analysis. The unit of analysis was the first ICU stay, with the time origin at ICU admission; stays with fewer than three charted motor examinations were excluded.

### Data Sources and Variables

Overt command-following was the Glasgow Coma Scale motor response charted by bedside nurses as ”Obeys Commands” (chartevents item 223901, score 6); any lower value was non-following (value set, eTable 2). The endpoint is the overt charted response, not a standardized disorders-of-consciousness diagnosis, and can be depressed by aphasia, motor-pathway injury, sedation, encephalopathy, or fluctuating arousal as well as a depressed level of consciousness; it cannot detect covert consciousness. A baseline non-follower had a first motor examination below threshold within 12 hours of ICU admission (70 patients [1.3%] without an examination in that window were classified from their first available examination).

Continuous infusions were resolved by drug class, separating hypnotic sedatives (propofol, midazolam, dexmedetomidine, ketamine, lorazepam, pentobarbital) from opioid analgesia (fentanyl), neuromuscular blockers, and vasopressors. Each examination was tagged for active infusion of each class, concurrent invasive ventilation, and, where a Richmond Agitation-Sedation Scale (RASS) value was charted within 2 hours (76.5% of examinations; item 228096), sedation depth, with RASS -4 or -5 defining deep sedation. Examinations during active neuromuscular blockade (0.87%; eTable 10) were carried forward at the last interpretable value; excluding them changed no estimate by more than one percentage point. Mechanical ventilation, demographics, discharge disposition, and in-hospital mortality were extracted from the procedure, admission, and patient tables.

### Outcomes and Measures

The primary measure was the time from ICU admission to first command-following among baseline non-followers, observed to ICU discharge and censored at 14 days, in a competing-risks framework[20] with three absorbing states: first command-following (the event of interest), in-hospital death or discharge to hospice, and alive ICU discharge without ever following commands; the last was modeled as its own competing event because it terminates observation and is clinically informative rather than non-informative censoring. Among patients who recovered at least once, instability was characterized by whether the first command-following was transient (the next examination fell below threshold) or sustained, the number of threshold crossings (also per 10 examinations and per ICU day), and whether recovery held through the final 24 hours. Sedation dependence was the proportion of non-following examinations during hypnotic sedation, opioid analgesia, and deep sedation (RASS -4 or -5), and the relative risk across each contrast. Discharge home served as a coarse functional-outcome proxy (no structured scale exists in the database) and was prespecified as exploratory.

Prespecified sensitivity analyses used a sustained-recovery endpoint (the final uninterrupted command-following period held to discharge), a two-consecutive-examination definition, strata of the baseline motor pattern, and a re-derivation using only interpretable examinations (free of neuromuscular blockade, deep sedation, and active hypnotic infusion); the competing death-or-comfort-care event was also separated into death and hospice. Full definitions are in the eMethods.

### Statistical Analysis

Cumulative incidence functions for command-following and for the competing death-or-comfort-care pathway were estimated with the Aalen-Johansen estimator, overall and by phenotype, and reported at 24 hours, 72 hours, 7 days, and 14 days with percentile bootstrap 95% confidence intervals (400 resamples). Equality across phenotypes was tested with a Gray-type permutation test on the integrated absolute difference between curves (1,000 permutations), with pairwise contrasts against ischemic stroke at 72 hours, and the restricted mean time in the command-following state over 14 days was estimated by phenotype. Adjusted associations with recovery timing were estimated with both a cause-specific Cox proportional-hazards model and a Fine-Gray subdistribution-hazard model (treating death or hospice and alive discharge without recovery as competing events), each adjusted for phenotype (ischemic stroke reference), age, sex, baseline sedation, and mechanical ventilation; a cause-specific Cox model for the competing death event used the same covariates. The proportional-hazards assumption (scaled Schoenfeld residuals) was not met for phenotype, age, and ventilation, so hazard ratios are interpreted as time-averaged and the non-parametric cumulative incidence functions are the primary estimand. Resampling was at the hospitalization level. Because 6.3% of admissions were repeat admissions from the same patient, the principal instability estimates were re-derived restricting to one stay per patient and with bootstrap and model-based 95% confidence intervals clustered on the patient. RASS values, used only for sedation-depth stratification, were missing for 23.5% of examinations and were not imputed; the deep-sedation contrast was additionally bounded under the extreme assumptions that every examination with a missing score was, or was not, deeply sedated (eMethods).

Transient-recovery proportions were compared with the chi-square test (Cramer V); threshold-crossing counts with the Kruskal-Wallis test (epsilon-squared) and negative-binomial regression carrying an examination-count offset (and, separately, an ICU-day offset), so incidence-rate ratios describe the per-examination rate. The association between crossings and discharge home was estimated with logistic regression as an exploratory analysis (odds ratio per crossing; age-and-phenotype-adjusted and fully adjusted models in eTable 14). Non-following relative risks used a stay-clustered bootstrap (2,000 resamples). The primary phenotype and instability family was corrected with the Benjamini-Hochberg false discovery rate; the threshold was P < .05, two-sided.

### Predictive Probes (Gradient Boosting, Deep Temporal Model, and a Local Language Model)

Three models probed how much of the eventual sustained command-following is encoded in the first 24 hours, among baseline non-followers surviving at least 24 hours: a gradient-boosted classifier over first-day tabular summaries, a gated recurrent unit network over the raw examination sequence, and a small open-weight large language model (Qwen3.5-4B, served locally through Ollama, 4-bit quantization, temperature 0) given the first-day flowsheet as text and asked, zero-shot, for the probability of sustained command-following, evaluated on a held-out random sample of 700 patients.[22] Models were evaluated by 5-fold stratified cross-validation (area under the receiver operating characteristic curve with bootstrap 95% confidence interval, Brier score, calibration, and decision-curve net benefit). To test whether the bedside fluctuation is structured or stochastic, the recurrent network was also trained to predict the next examination’s state and compared with a first-order Markov baseline. These were measurement probes, not deployable prognostic tools, and were not externally validated; model cards and feature lists are in the eMethods. Analyses used Python 3.9 (pandas, scikit-learn, statsmodels, SciPy, lifelines, PyTorch); code is at https://github.com/Alon-Gorenshtein/study_command_following.

### Ethics

MIMIC-IV contains de-identified data maintained under a data use agreement overseen by the institutional review boards of the Massachusetts Institute of Technology and Beth Israel Deaconess Medical Center, which granted a waiver of informed consent for the original collection; access was through credentialed PhysioNet authorization. This secondary analysis of a publicly available de-identified database required no additional institutional review board approval.

## Results

### Cohort

Of 94,458 ICU stays in MIMIC-IV, 14,272 were first ICU stays of adults with an acute brain injury, of whom 13,900 (97.4%) had three or more charted motor examinations and formed the analytic cohort (Figure 1). The median age was 67 years (IQR, 54 to 78), 44.2% were women, 37.8% were mechanically ventilated, and 43.9% received continuous sedation or analgesia. In-hospital mortality was 17.9% and 31.5% of patients were discharged home. At admission, 5,498 patients (39.6%) were not following commands; this proportion ranged from 30.4% in subdural hemorrhage to 59.4% in anoxic injury (Table 1).

**Figure 1.**
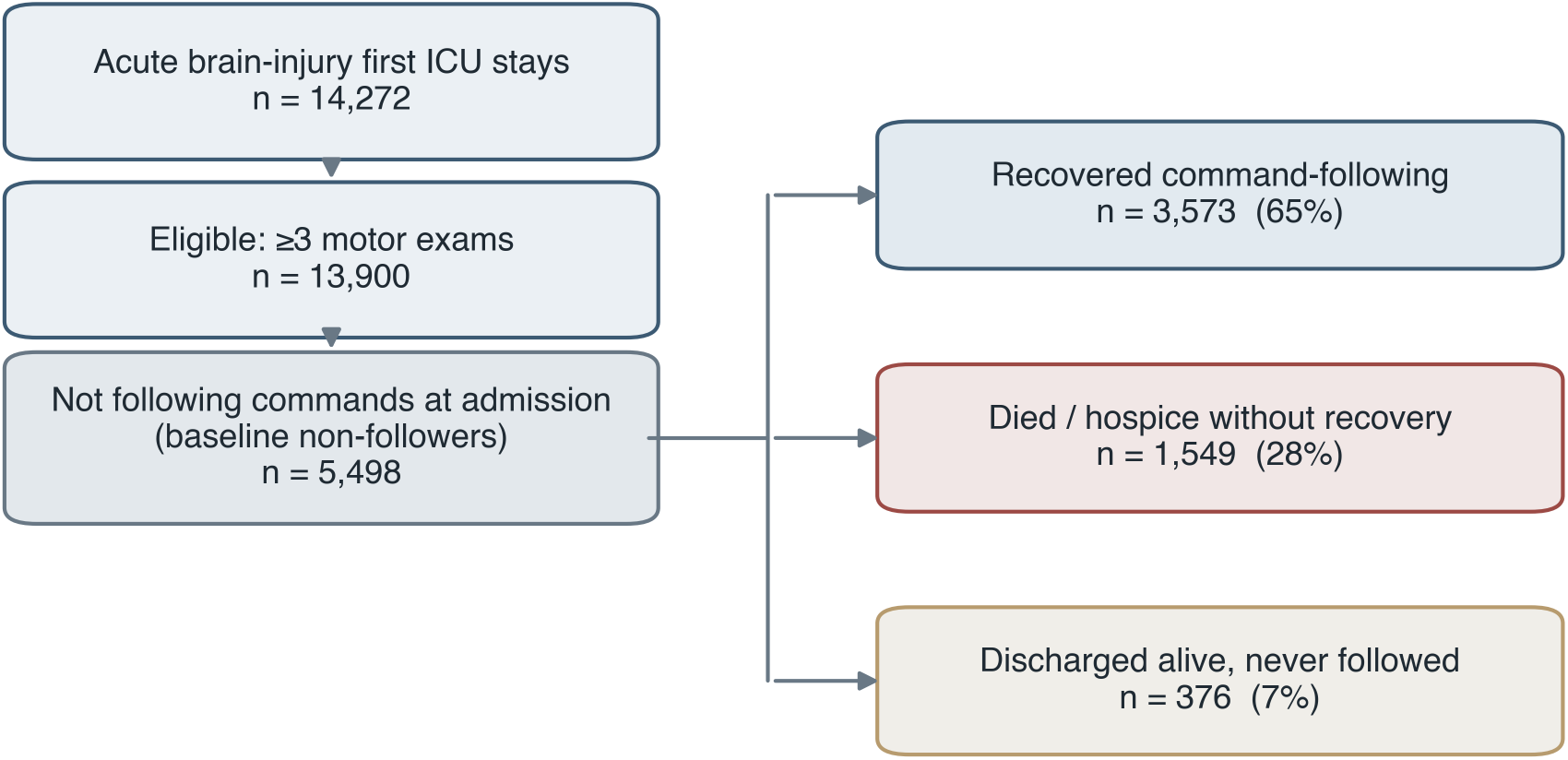
Study flow. Derivation of the analytic cohort and baseline non-followers, and their resolution into recovery of command-following, death or hospice, and alive discharge without command-following.

**Table 1.** Cohort characteristics by acute brain-injury phenotype. AIS, acute ischemic stroke; TBI, traumatic brain injury; ICH, intracerebral hemorrhage; SAH, subarachnoid hemor-rhage; SDH, subdural hemorrhage; Anoxic, anoxic-ischemic injury.

| Characteristic | Anoxic | AIS | TBI | SDH | ICH | SAH | Overall |
| --- | --- | --- | --- | --- | --- | --- | --- |
| No. of patients | 1244 | 4021 | 3787 | 780 | 2784 | 1284 | 13900 |
| Age, median (IQR), y | 59<br>(48-69) | 70<br>(60-80) | 65<br>(47-80) | 70<br>(59-79) | 69<br>(57-80) | 61 (50-72) | 67<br>(54-78) |
| Female, % | 43.6 | 48.0 | 37.3 | 36.8 | 45.2 | 56.2 | 44.2 |
| Mechanically ventilated, % | 55.1 | 34.5 | 37.3 | 29.1 | 35.2 | 44.0 | 37.8 |
| Any continuous sedation or analgesia, % | 57.3 | 41.6 | 45.1 | 34.4 | 39.2 | 50.2 | 43.9 |
| Motor exams, median (IQR), No. | 24<br>(14-45) | 27<br>(13-48) | 23<br>(11-48) | 28<br>(15-52) | 37<br>(18-77) | 76<br>(29-155) | 29<br>(14-60) |
| Not following at admission, % | 59.4 | 36.7 | 37.5 | 30.4 | 39.3 | 41.5 | 39.6 |
| In-hospital mortality, % | 39.9 | 14.8 | 11.1 | 15.0 | 20.8 | 21.8 | 17.9 |
| Discharged home, % | 27.0 | 31.6 | 37.9 | 33.7 | 21.7 | 36.4 | 31.5 |

### Recovery of Command-Following (Aim 1)

#### Command-following returned in most baseline non-followers, early and at a phenotype-dependent rate

Among the 5,498 patients not following commands at admission, the three-state competing-risks cumulative incidence of first command-following was 43.5% (95% CI, 42.4 to 44.9) by 24 hours, 55.1% (54.0 to 56.7) by 72 hours, 61.4% (60.2 to 62.6) by 7 days, and 65.0% (64.0 to 66.3) by 14 days, with a median of 12.2 hours (Figure 2); by 14 days the competing incidence of death or hospice was 28.2% and that of alive discharge without recovery 6.8%. Recovery was frequently not maintained: sustained command-following (the final uninterrupted following period held to discharge) reached only 52.1% by 14 days, 13 percentage points below the 65.0% who followed at least once, and took a median of 32 hours (Figure 2A, dashed line; eTable 15). The 14-day incidence ranged more than twofold across phenotypes, from 36.9% in anoxic injury and 58.3% in subarachnoid hemorrhage to 73.8% in traumatic brain injury and 77.2% in ischemic stroke (Figure 2). Relative to ischemic stroke, the 72-hour incidence was lower in anoxic injury (difference, 0.41; adjusted P = .002), subarachnoid hemorrhage (0.17; P = .002), and intracerebral hemorrhage (0.16; P = .002), modestly lower in subdural hemorrhage (0.07; unadjusted P = .04), and did not differ in traumatic brain injury (0.02; P = .28); the curves differed overall (Gray-type permutation test, P = .001). The restricted mean time having achieved command-following over 14 days ranged from 4.2 days (95% CI, 3.8 to 4.6) in anoxic injury to 9.6 days (9.3 to 9.9) in ischemic stroke. The gradient persisted after adjustment for age, sex, baseline sedation, and ventilation: the Fine-Gray subdistribution hazard relative to ischemic stroke was 0.41 (95% CI, 0.37 to 0.46) in anoxic injury, 0.72 (0.64 to 0.81) in subarachnoid hemorrhage, and 0.75 (0.69 to 0.82) in intracerebral hemorrhage, with concordant cause-specific hazard ratios (Figure 3; eTable 7). Baseline sedation carried a higher subdistribution hazard of recovery (1.46; 95% CI, 1.36 to 1.57), consistent with reversible pharmacological suppression. Estimates were stable across sensitivity analyses (excluding neuromuscular-blockade examinations, 72-hour incidence 55.8% and transient recovery 22.1%; single-phenotype hospitalizations, 57.6%; eTables 12 and 13). Recovery varied with the baseline motor pattern: the 14-day incidence ranged from 79.6% in patients who localized to pain and 66.5% with flexion-withdrawal to 34.6% with abnormal flexion and 29.9% with abnormal extension, and patients with no motor response recovered more often (58.6%) than those with posturing (eTable 16). The competing event was almost entirely in-hospital death (1,867 patients) rather than hospice (212; 1.9% by 14 days), so recovery estimates were not materially shaped by early withdrawal-of-care transitions.

**Figure 2.**
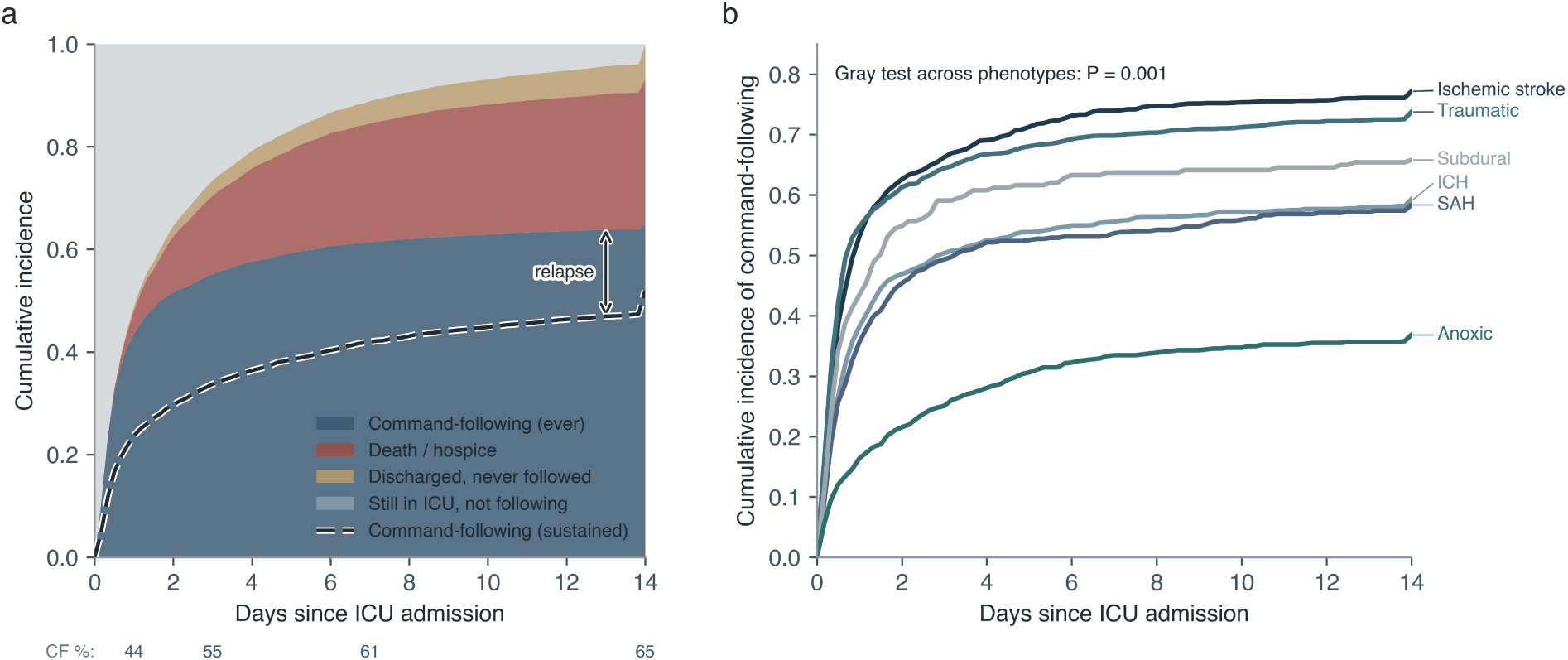
Competing-risks recovery of command-following. (a) Stacked cumulative incidence of first command-following, the competing death-or-hospice pathway, alive discharge without recovery, and remaining in the ICU, over 14 days among baseline non-followers, with the bootstrap 95% confidence band; the dashed line is sustained command-following (the final following period held to discharge), and the gap between it and the top of the command-following band is relapse. (b) Command-following cumulative incidence by acute brain-injury phenotype (Gray-type permutation test across phenotypes, P = .001).

**Figure 3.**
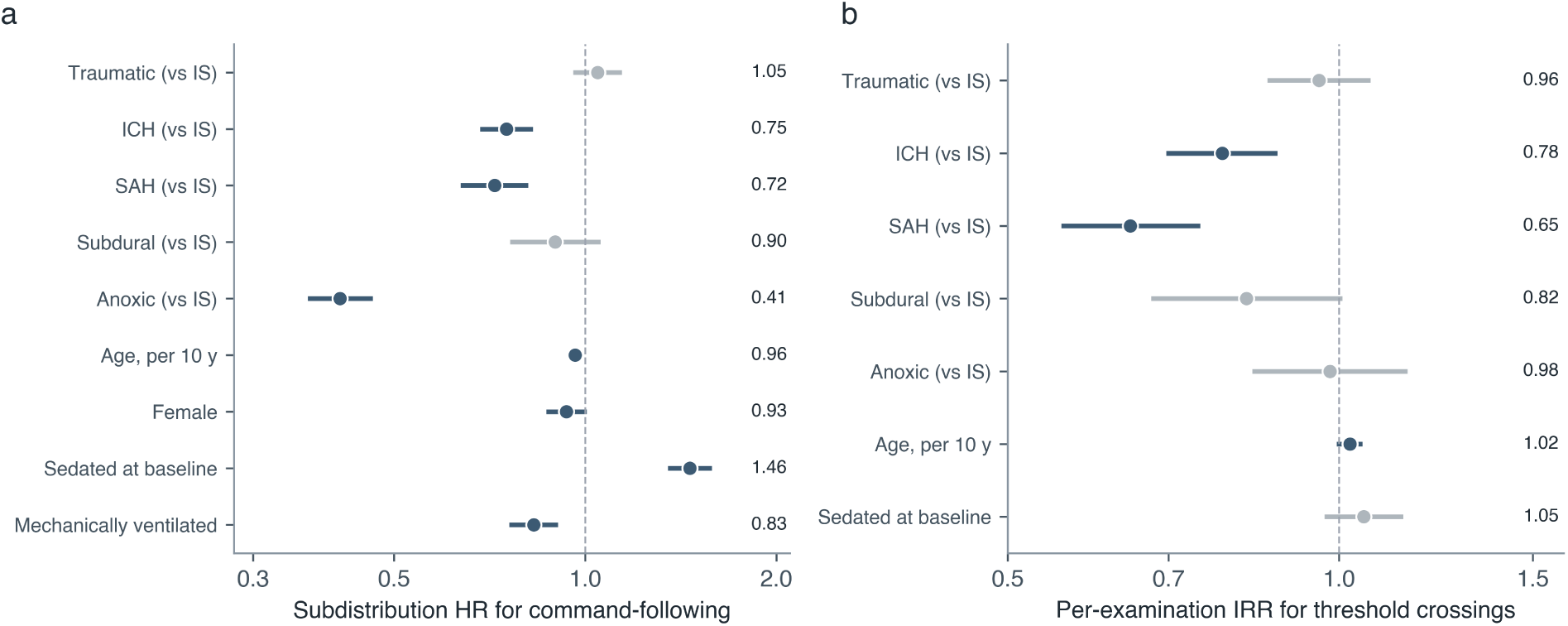
Adjusted determinants. (a) Fine-Gray subdistribution hazard ratios for command-following, adjusted for phenotype, age, sex, baseline sedation, and mechanical ventilation (ischemic stroke reference). (b) Negative-binomial incidence-rate ratios for the number of threshold crossings. Grey denotes non-significant terms.

### Instability of Recovery (Aim 2)

#### Once command-following appeared, it remained unreliable

Among examinations after a patient’s first command-following, command-following was present at only 76% in the first 6 hours, declined to about 60% over the following week, and was confirmed at the immediately following examination in only 77.8%; more than a third of post-recovery examinations did not show command-following (eFigure 1). Of the 3,573 patients who recovered, only 37.6% did so as a single transition that held to discharge, 42.4% relapsed at least once before following at discharge, and 20.0% were not following at discharge despite having followed earlier. The first recovery was transient (a fall below threshold at the next examination) in 22.2% (95% CI, 20.9 to 23.6), and 62.4% (60.8 to 64.0) crossed the threshold in and out at least once, with a median of 3 crossings (mean, 5.0). The transient-recovery proportion did not differ across phenotypes (Cramer V, 0.04; adjusted P = .32). The higher raw crossing count in the hemorrhagic phenotypes was largely an artifact of examination frequency: after an examination-count offset, the per-examination rate was lower in subarachnoid hemorrhage (incidence-rate ratio, 0.65; 95% CI, 0.56 to 0.74) and intracerebral hemorrhage (0.78; 0.70 to 0.88) than in ischemic stroke, and per 10 examinations the median rate was similar across phenotypes (0.51 to 0.86). Instability was therefore a general property that accumulates with surveillance. In an exploratory model, more crossings were associated with lower odds of discharge home after full adjustment for age, phenotype, vasopressor use, sedation exposure, length of stay, and ventilation (odds ratio per crossing, 0.90; 95% CI, 0.87 to 0.94).

### The Sedation Component (Aim 3)

#### Non-following depended strongly on sedation, and most strongly on its depth

Among examinations with a concurrent RASS value, 98.0% were non-following when the patient was deeply sedated (-4 or -5) versus 28.1% otherwise (relative risk, 3.49; 95% CI, 3.37 to 3.61). Active hypnotic sedation carried a higher risk than opioid analgesia (relative risk, 2.36; 95% CI, 2.28 to 2.45, versus 2.18; 2.11 to 2.26), and examinations during invasive ventilation were non-following far more often (74.0% versus 19.8%). The dependence persisted beyond the infusion: examinations within 6 hours of stopping a hypnotic were non-following more often than those long off sedation (51.8% versus 41.0%; Figure 4; eTable 9). Two subgroups defined by sedation practice recovered differently: patients never deeply sedated reached command-following by 72 hours far more often than the cohort (75.0% versus 55.1%), whereas the 1,702 patients who never received any continuous sedative or analgesic recovered less often (47.5%); this is a selected population whose non-following more often reflected structural injury, so these contrasts are descriptive. Re-estimating recovery using only interpretable examinations (free of blockade, deep sedation, or active hypnotic infusion) lowered the 72-hour incidence from 55.1% to 48.3% while leaving the 14-day incidence essentially unchanged (65.3%), so part of the apparently rapid early recovery reflects the lifting of sedation rather than neurological change (eTable 15). The principal estimates were robust in sensitivity analyses: restricting to one stay per patient left the 14-day cumulative incidence (65.7% versus 65.0%) and the transient-recovery proportion (22.4% versus 22.2%) essentially unchanged, and 95% confidence intervals clustered on the patient rather than the stay were materially unchanged and continued to exclude the null. The deep-sedation association persisted under the extreme assumptions that every examination with a missing RASS was deeply sedated (relative risk, 2.04) or was not (3.21), bounding the available-case estimate (eMethods).

**Figure 4.**
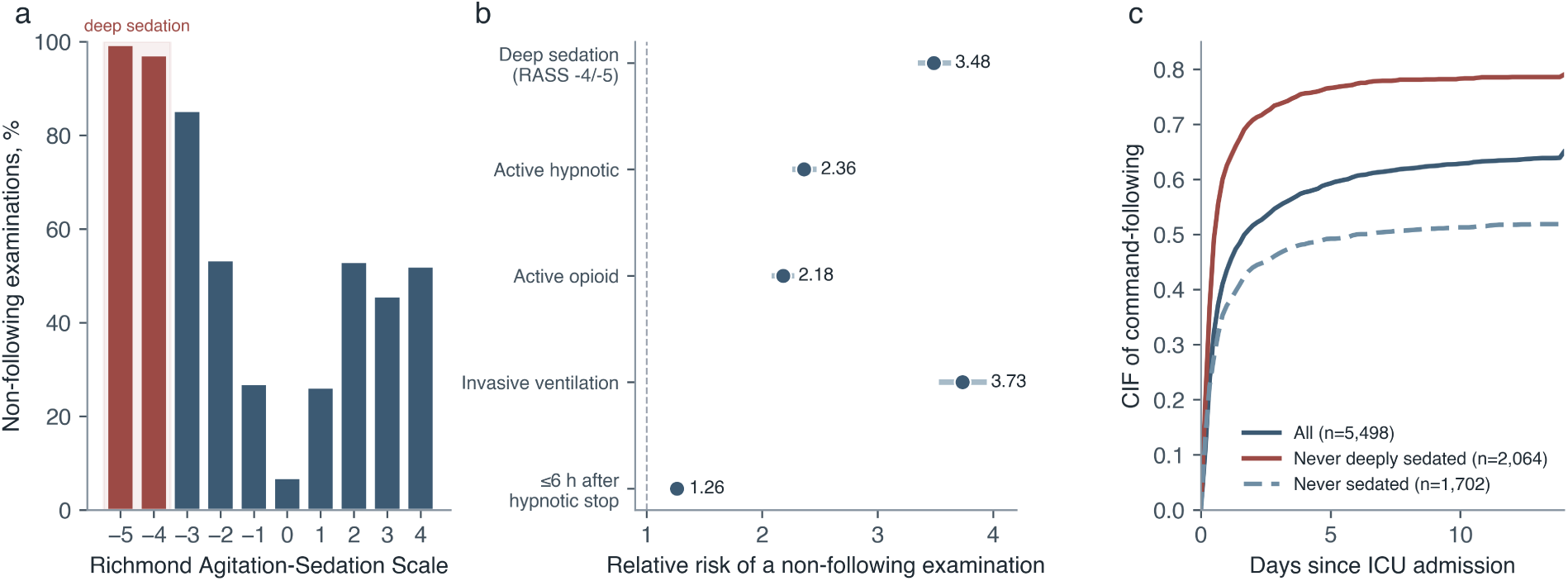
Sedation depth and non-following examinations. (a) Percentage of examinations that were non-following by Richmond Agitation-Sedation Scale value; deep sedation (-4/-5) is shaded. (b) Relative risk of a non-following examination for each sedation contrast, with 95% confidence intervals. (c) Three-state cumulative incidence of command-following in all baseline non-followers, in those never deeply sedated, and in those never given any continuous sedative or analgesic.

### Predictability of Recovery and of Fluctuation (Aim 4)

#### The first day of bedside behavior carried substantial but incomplete information, and the fluctuation itself was highly structured

In a measurement probe (eMethods, eTable 8), a gradient-boosted model using only first-day features discriminated sustained recovery with an area under the receiver operating characteristic curve of 0.82 (95% CI, 0.81 to 0.83), well above an age-and-phenotype baseline (0.64) but leaving many patients undetermined; a deep temporal model did not improve on it, and a small zero-shot language model reached 0.75, so the early signal is informative but shallow. The bedside state was also highly persistent between consecutive examinations, so the fluctuation accumulating across a stay arose from a small per-examination probability of change rather than from disorganized noise; the next-examination forecasting analysis and its first-order Markov comparison are reported in the eMethods (eFigure 2).

### External Validation in a Multicenter Cohort

#### The recovery trajectory and its instability reproduced across 208 hospitals

We repeated the analysis in the eICU Collaborative Research Database (eICU-CRD v2.0; 208 U.S. hospitals, 2014 to 2015), holding the command-following construct, the six ICD-based phenotypes, and the competing-risks and instability estimators identical (eMethods S1.9; eTable 17). Of 12,397 first ICU stays of adults with an acute brain injury, 7,615 had three or more charted Glasgow Coma Scale motor examinations, and 2,519 were not following commands at admission. The cumulative incidence of first command-following was 64.8% by 14 days (95% CI, 62.7 to 67.0), closely matching the 65.0% observed in MIMIC-IV, with a median of 16.2 hours; early incidence was lower (35.1% at 24 hours, 51.3% at 72 hours) and converged by 7 to 14 days, consistent with sparser first-day motor charting and shorter unit stays in eICU. The phenotype gradient was preserved, from 29.1% at 14 days in anoxic injury to 79.3% in ischemic stroke, in the same rank order as the primary cohort. The instability of recovery also replicated: among the 1,479 patients who recovered, the first return of command-following was transient in 22.7% (95% CI, 20.6 to 24.9), against 22.2% in MIMIC-IV; 64.2% crossed the threshold at least once after recovering (MIMIC-IV, 62.4%); and the median number of crossings was again 3. The principal findings, early phenotype-dependent recovery and its subsequent instability, therefore reproduced in an independent multicenter cohort.

## Discussion

In a cohort of 14,272 brain-injured ICU patients, overt bedside command-following returned in about two thirds of those unresponsive at admission, usually within the first days and at a rate that differed more than twofold across phenotypes. The central finding is not that recovery occurs but that it is unsteady: the first return was lost at the next examination in more than one in five patients, and most patients crossed the responsiveness threshold repeatedly rather than once. This instability was a general property of recovery rather than a feature of any phenotype; the larger raw crossing count in the hemorrhagic phenotypes reflected their more frequent examination, and per examination the phenotypes were similar. Non-following depended strongly on sedation, most strongly on its depth, with nearly all deeply sedated examinations showing no command-following. Overt bedside command-following is therefore an early but unstable and sedation-sensitive signal whose single observations should not be read as fixed evidence about the underlying state. That responsiveness fluctuates and that sedation suppresses the examination are recognized qualitatively in specialist cohorts; this work quantifies these properties, and the gap between ever following commands and following them durably, at the scale of routine acute intensive care.

These findings extend to the acute ICU a property of consciousness documented mainly in rehabilitation cohorts. Fluctuation of responsiveness is a defining feature of the minimally conscious state and a recognized cause of bedside diagnostic error, where serial structured assessment is recommended because a single examination is unreliable.[6,7,16] The present analysis quantifies that instability at the scale of routine care and shows it begins in the first hours and days after injury, not only in the chronic phase. The phenotype gradient, lowest and slowest in anoxic injury and highest in ischemic stroke and traumatic brain injury, is consistent with the known prognostic ordering of these injuries and supports the face validity of the bedside milestone as a coarse marker of trajectory.[5,10,17]

The work also clarifies how sedation shapes the examination. Measuring its footprint by depth and class showed a graded association: nearly all deeply sedated examinations and three quarters of examinations during invasive ventilation showed no command-following, hypnotics carried more risk than opioids, and a residual effect persisted in the hours after a hypnotic was stopped. The charted examination thus reflects two processes, reversible pharmacological suppression and the slower resolution of injury, and an electronic-record command-following endpoint that ignores sedation will misattribute the timing of recovery. We did not attempt a causal decomposition: the two sedation-practice subgroups recovered in opposite directions (never-deeply-sedated faster, never-sedated slower), showing they mark different populations. Consistent with a reversible component, baseline sedation carried a higher subdistribution hazard of recovery.

The predictive probes were measurements, not proposed tools, and what they did not show is as informative as what they did. First-day bedside dynamics predicted sustained recovery well above an age-and-phenotype baseline, yet many patients remained undetermined on day 1, so the early examination is informative but far from sufficient. A deep temporal model over the raw sequence did not improve on simple first-day summaries, and a small zero-shot language model reading the flowsheet text approached but did not exceed them; complex temporal and language models added little, and the value of the examination lies in its repetition. Because the probes were restricted to first-day survivors and used a discharge-anchored label, their discrimination partly reflects the observation window and is not a prognostic claim. The forecasting analysis showed the bedside state is highly persistent between examinations, so the fluctuation accumulating across a stay arises from a small per-examination probability of change, a structured property of the recovering brain rather than measurement noise.

This study has several limitations. First, the endpoint is overt, charted command-following, not a standardized disorders-of-consciousness diagnosis; non-following can arise from aphasia, motor-pathway injury, sedation, or encephalopathy as well as depressed consciousness, and covert consciousness and cognitive-motor dissociation are undetectable without task-based electroencephalography or functional imaging, so the non-follower group certainly includes aware patients.[18,19] That the overt signal is unstable and sedation-sensitive strengthens the case for systematic advanced assessment in this group. Second, the database has no validated functional outcome; discharge disposition is a coarse proxy, so the fluctuation–discharge-home association is exploratory. Third, both the timing of recovery and the crossing count depend on examination frequency; we addressed this with an examination-frequency floor and by expressing crossings per examination and per ICU day, but charting intensity and true fluctuation cannot be fully separated in routine data. Because the first command-following is a single nurse-charted observation, an isolated reflexive ”Obeys Commands” would inflate the transient-recovery and crossing rates, though the conservative two-consecutive-examination endpoint (eTable 15) showed the same pattern; and because the state is observed only at charted examinations, the reported median times are upper bounds. Fourth, the sedation-practice subgroups are selected populations, so the sedation contrasts are descriptive, and the deep-sedation contrast partly reflects a floor effect of arousal on the examination (command-following is near-impossible at RASS -4 or -5). Fifth, the proportional-hazards assumption failed for some covariates, which is why the cumulative incidence functions are the primary estimand. Sixth, withdrawal of life-sustaining therapy both competes with and influences the observation of recovery; the competing-risks design handles the former but not the latter. Seventh, phenotypes were defined from administrative codes in any position rather than chart adjudication, admitting misclassification; the concordant single-phenotype sensitivity analysis bounds this concern. Finally, the primary cohort is single-center (2008 to 2019); we externally validated the recovery trajectory, its phenotype gradient, and its instability in an independent multicenter cohort of 208 hospitals (eICU-CRD), where the 14-day incidence (64.8%) and transient-recovery rate (22.7%) closely matched, although both sources are U.S. intensive-care electronic records, and a prospective comparison against standardized serial assessment remains needed.

These findings should be read against the standardized disorders-of-consciousness assessments they do not replace. The Glasgow Coma Scale motor score is coarser than the Coma Recovery Scale-Revised, which is more sensitive to covert awareness but hard to administer serially in unstable patients; intensive-care-oriented instruments such as the CRSR-FAST and the Simplified Evaluation of Consciousness Disorders were developed to make standardized assessment feasible at the bedside.[21,16] That repeated standardized assessment reduces misdiagnosis is established,[7] and these results extend that principle to the acute phase using data already collected. Prospective work should compare electronic-record command-following trajectories against serial standardized assessments and against electroencephalographic or functional-imaging markers of covert command-following.

For clinical practice, the results argue against treating any single charted observation of command-following as a stable indicator of the underlying state, and in favor of serial, sedation-aware assessment interpreted against sedation depth before that milestone anchors prognosis. For research, a command-following endpoint derived from the electronic record should be defined over repeated examinations, account for sedation depth, and be normalized for examination frequency when fluctuation is compared across groups. Future work should link this instability to validated functional outcomes and to quantitative measures of arousal and covert command-following.

## Conclusion

After acute brain injury, overt bedside command-following returns early but unsteadily, with phenotype-dependent timing, frequent fluctuation across the responsiveness threshold, and a strong dependence on sedation depth. The findings concern the overt, charted examination used in routine ICU care, not consciousness itself, and within that scope they indicate that a single observation is an unreliable index and that command-following is better measured as a fluctuating process than as a single event.

## Supporting information

Full Appendix

## Data Availability

All data produced are available online at https://github.com/Alon-Gorenshtein/study_command_following

## Declarations

### Author contributions

Contributions are described using the CRediT taxonomy. A.G.: conceptualization, methodology, software, formal analysis, data curation, validation, visualization, and writing of the original draft. Y.A.: software, data curation, formal analysis, validation, and review and editing of the manuscript. M.O.: methodology, software, validation, and review and editing of the manuscript. Y.B.: methodology, validation, and review and editing of the manuscript. E.K.: conceptualization, methodology, supervision, resources, and review and editing of the manuscript. O.D.: conceptualization, clinical interpretation, supervision, and review and editing of the manuscript. All authors critically reviewed the manuscript and approved the final version submitted for publication. A.G. (corresponding author) had full access to all data in the study and takes responsibility for the integrity of the data and the accuracy of the analysis.

### Originality and prior publication

This manuscript describes original work. It has not been published previously, in whole or in part, and is not under consideration for publication by any other journal. All authors have read and approved the submitted manuscript.

### Ethics approval and informed consent

This was a retrospective analysis of de-identified data from two publicly available critical care databases: MIMIC-IV (version 3.1) and the eICU Collaborative Research Database (version 2.0), each accessed under a credentialed PhysioNet data use agreement. The original collection of MIMIC-IV was approved by the institutional review boards of the Massachusetts Institute of Technology and Beth Israel Deaconess Medical Center, which granted a waiver of informed consent; the eICU Collaborative Research Database was likewise approved with a waiver of informed consent for the use of de-identified data. Because the present study used only previously collected, fully de-identified records and involved no patient contact and no intervention, it qualified as non-human-subjects research and required no additional institutional review board approval or informed consent at the authors’ institutions. The study was conducted in accordance with the principles of the Declaration of Helsinki.

### Reporting checklist

The study was reported in accordance with the STROBE (Strengthening the Reporting of Observational Studies in Epidemiology) statement, with the RECORD (REporting of studies Conducted using Observational Routinely collected health Data) extension for studies that use routinely collected health data. The prediction-model component was additionally reported following the TRIPOD (Transparent Reporting of a multivariable prediction model for Individual Prognosis Or Diagnosis) statement. The completed STROBE checklist with RECORD items is uploaded as a separate file with this submission.

### Use of artificial intelligence

A large language model was used solely to assist with grammar and language editing. It was not used for any other purpose. The authors reviewed all text and take full responsibility for the content of the manuscript.

