## Supplementary material for "The Unsteady Return of Command-Following: Recovery and Instability of Bedside Motor Command-Following After Acute Brain Injury": Full Appendix

**Affiliations:**

This Supplementary Information accompanies the main manuscript. It contains the operational cohort definitions and diagnosis code lists, the full statistical detail for the competing-risks and instability analyses, the sedation-tagging procedure, the measurement-probe model card, and supporting tables and figures. All values are reproduced from the analysis pipeline; no individual-level data are included.

**Contents**


- S1 Supplementary Methods
- S2 Supplementary Tables (eTable 1 to eTable 17)
- S3 Supplementary Figures (eFigure 1 to eFigure 2)
- S4 Code and Data Availability


### S1. Supplementary Methods

#### S1.1 Data source and cohort

Data came from the Medical Information Mart for Intensive Care IV (MIMIC-IV), version 3.1, a single-center critical care database of adults admitted to the intensive care units of Beth Israel Deaconess Medical Center between 2008 and 2019. Adults (age 18 years or older) with an acute brain injury diagnosis recorded for the hospitalization were eligible. The first intensive care unit (ICU) stay of each hospitalization was used so that observations were not clustered within a hospitalization across ICU transfers. Stays with fewer than three charted Glasgow Coma Scale motor examinations were excluded so that a within-stay trajectory could be observed; this requirement retained 13,900 of 14,272 brain-injury first stays (97.4%).

#### S1.2 Phenotype assignment

Each hospitalization was assigned to one of six acute brain-injury phenotypes from International Classification of Diseases, Ninth and Tenth Revision (ICD-9 and ICD-10) diagnosis codes (eTable 1). Codes were matched in any diagnosis position (a hospitalization qualified if any of its recorded diagnoses matched a phenotype code list), not only the principal diagnosis. This any-position rule was chosen to maximize sensitivity, because the brain injury is frequently not the principal diagnosis (anoxic injury, for example, is usually coded secondary to a cardiac-arrest principal diagnosis); the cost is reduced specificity, since a matching code in a non-principal position need not be the index event of the admission, may reflect a historical or resolved condition, and was not adjudicated against the chart. When more than one phenotype was present, a single primary phenotype was assigned by a fixed clinical priority hierarchy, in which the more procedurally specific and acute diagnosis took precedence: subarachnoid hemorrhage, then intracerebral hemorrhage, subdural hemorrhage, traumatic brain injury, ischemic stroke, and anoxic injury.

##### S1.3 The command-following measure

Command-following was operationalized as the Glasgow Coma Scale motor response recorded by bedside nurses as "Obeys Commands" (chartevents item 223901, mapped to a motor score of 6). A motor response below this value was classified as non-following. The full enumerated value set and its numeric mapping are given in eTable 2. A patient was classified as a baseline non-follower when the first motor examination within 12 hours of ICU admission was below the command-following threshold.

##### S1.4 Competing-risks cumulative incidence

Among baseline non-followers, the time from ICU admission to the first command-following examination was analyzed as a three-state competing-risks problem with three mutually exclusive absorbing states: first command-following (the event of interest), in-hospital death or discharge to hospice (the death or comfort-care pathway), and alive ICU discharge without ever following commands. Alive discharge without recovery was modeled as its own competing event rather than as administrative censoring, because it terminates observation and is clinically informative; observation was otherwise censored at 14 days. Cumulative incidence functions were estimated with the Aalen-Johansen estimator, computed in-house from the cause-specific event and at-risk counts at each event time, and were evaluated at 24 hours, 72 hours, 7 days (168 hours), and 14 days (336 hours); at 14 days the three cumulative incidences summed to the cohort. Percentile 95% confidence intervals were obtained from 400 bootstrap resamples of admissions (the unit of analysis was the brain-injury hospitalization, taken as its first ICU stay). Between-phenotype differences in the command-following cumulative incidence at 72 hours were tested against the ischemic stroke reference with a two-sided permutation test (1,000 permutations of the phenotype label).

##### S1.5 Instability metrics

Instability of recovery was characterized among patients who reached command-following at least once. Three per-patient measures were computed from the ordered motor trajectory. A first recovery was transient when the examination immediately following the first "Obeys Commands" fell below the threshold, and sustained otherwise. The number of threshold crossings counted every transition of the binary follow or not-follow indicator across the stay, so that a patient who rose to command-following and never fell had one crossing, and a patient who moved in and out repeatedly had several. A recovery was held when every examination in the final 24 hours of the trajectory was at the command-following threshold. Because the number of crossings depends on examination frequency, it was modeled with negative-binomial regression carrying an offset for the number of examinations (and separately for ICU days), and was also expressed per 10 examinations and per ICU day. The exploratory association between the number of crossings and discharge home (home or home with services) was estimated with logistic regression; we report a model adjusted for age and phenotype and a more fully adjusted model that also includes vasopressor use, sedation exposure, length of stay, and mechanical ventilation (eTable 14). Because discharge disposition is shaped by social and systemic factors not captured here, this analysis is exploratory and is not a prognostic claim.

##### S1.6 Sedation tagging and decomposition

Continuous infusions were extracted from the infusion records as timestamped start-to-end intervals and resolved by drug class, separating hypnotic sedatives (propofol, midazolam, dexmedetomidine, ketamine, lorazepam, pentobarbital) from opioid analgesia (fentanyl), and from neuromuscular blocking agents and vasopressors. Each motor examination was tagged for active infusion of each class, for concurrent invasive ventilation, for sedation depth where a Richmond Agitation-Sedation Scale value was charted within 2 hours (deep sedation defined as -4 or -5), and for time since the last hypnotic was stopped. Because neuromuscular blockade abolishes the motor response, examinations recorded during active blockade were carried forward at the last interpretable value in the primary analysis; excluding them entirely was a sensitivity analysis that changed no estimate by more than one percentage point (eTable 10). The relative risk of a non-following result was estimated for each sedation contrast (deep versus light sedation, active hypnotic, active opioid, invasive ventilation, and the early post-discontinuation window) with a stay-clustered bootstrap (2,000 resamples of stays); each contrast, its denominators, and its reference group are given in eTable 9. The command-following cumulative incidence was re-estimated in the subset of baseline non-followers who were never deeply sedated and in the subset who never received any continuous sedative or analgesic infusion, as sensitivity analyses for the sedation contribution (Figure 4).

##### S1.7 Predictive probes

Three models asked, as a measurement question, how much of the eventual sustained command-following is encoded in the first 24 hours of bedside behavior, among baseline non-followers who survived at least 24 hours. The first was a gradient-boosted decision-tree classifier over tabular summaries of the first day (eTable 6). The second was a deep temporal model, a single-layer gated recurrent unit network (hidden size 48) implemented in PyTorch on the Apple Metal backend, which read the raw ordered sequence of first-day examinations, each step encoding the motor score, a command-following indicator, the sedation and neuromuscular-blockade flags, and the elapsed and inter-examination times; it was trained with binary cross-entropy and the Adam optimizer and evaluated by the same 5-fold cross-validation as the gradient-boosted model. The third was a small open-weight large language model (Qwen3.5-4B, the 4-billion-parameter dense variant of the Apache-2.0 Qwen3.5 release; served locally through Ollama as tag qwen3.5:4b, 4-bit quantization, temperature 0, thinking mode disabled, with answers constrained to an integer probability), which received each patient's first-day examination flowsheet rendered as plain clinical text and was asked, zero-shot and with no fine-tuning, for the probability of sustained command-following; it was evaluated on a held-out random sample of 700 patients and compared with the other models restricted to those same patients.

To test whether the bedside fluctuation is structured or stochastic, the recurrent network was also trained to predict, at each examination, the command-following state of the next examination from the sequence so far, and was compared with a first-order Markov baseline that used only the empirical probability of the next state given the current state. All three probes are descriptive: they quantify the information content of early bedside dynamics, are not deployable prognostic tools, were not calibrated for individual decision-making, and were not externally validated.

##### S1.8 Statistical transparency, missing data, and software

The Aalen-Johansen cumulative incidence estimator, the percentile bootstrap confidence intervals, and the Gray-type permutation test were implemented in-house in NumPy. Cause-specific Cox and Fine-Gray models used lifelines 0.30; the Fine-Gray subdistribution model was fit as an inverse-probability-of-censoring-weighted Cox model on a start-stop counting-process expansion. Consistent with the descriptive three-state cumulative incidence functions, both death or hospice and alive ICU discharge without recovery were treated as competing events, so subjects experiencing either remained in the subdistribution risk set after their event with decaying weights; the censoring distribution was estimated by the Kaplan-Meier method from administrative censoring at 14 days only (subjects still in the ICU, or first following commands after day 14). The proportional-hazards assumption was tested with scaled Schoenfeld residuals (rank-transformed time) and was violated for phenotype, age, and ventilation; the hazard ratios are therefore time-averaged, and the non-parametric cumulative incidence functions are the primary descriptive estimand. The unit of analysis was the brain-injury hospitalization (operationalized as its first ICU stay), and all bootstrap and permutation resampling was clustered at that admission level. The analytic cohort of 13,900 admissions came from 13,023 unique patients: 773 patients (5.9%) contributed more than one brain-injury hospitalization (maximum 6), so 877 admissions (6.3%) were repeat admissions of an earlier patient; among the 5,498 baseline non-followers, only 124 patients (2.3%) had more than one admission. Because repeat brain-injury admissions are infrequent and each is a clinically distinct event, the primary resampling unit was the admission; within-patient correlation was nonetheless addressed in sensitivity analyses by re-deriving the principal estimates restricting to one stay per patient (the 14-day cumulative incidence of command-following moved from 65.0% to 65.7%, and the transient-recovery proportion from 22.2% to 22.4%) and by attaching bootstrap and model-based 95% confidence intervals clustered on the patient rather than the admission, which left the intervals materially unchanged and continued to exclude the null. The negative-binomial models for threshold crossings carried an offset for the number of examinations (or ICU days). Richmond Agitation-Sedation Scale values were available within 2 hours for 76.5% of examinations and were used only for the sedation-depth stratification; they were not imputed. Missingness was approximately non-differential with respect to the examination state (non-following in 38.7% of examinations with a charted score versus 37.3% of those without), and the deep-sedation contrast was bounded under the extreme assumptions that every examination with a missing score was deeply sedated (relative risk, 2.04) or was not (3.21), both well above unity. A first motor examination within 12 hours of admission was present for 99.5% of patients; the 70 patients (1.3%) without one were classified from their first available examination. Examinations during neuromuscular blockade (0.87%) were excluded in the primary analysis. The deep temporal model used PyTorch 2.8 on an Apple Metal (MPS) backend; the language model was Qwen3.5-4B (4-bit quantized) served locally through Ollama at temperature 0. The significance threshold was $P < .05$, two-sided, and the primary family of contrasts was corrected with the Benjamini-Hochberg false discovery rate procedure.

#### S1.9 External validation in the eICU Collaborative Research Database

The principal findings were tested for reproducibility in the eICU Collaborative Research Database (eICU-CRD) version 2.0, a multicenter critical-care database of more than 200,000 ICU unit stays at 208 U.S. hospitals (2014 to 2015), accessed through PhysioNet. The analysis pipeline was transferred to eICU without changing the construct. The unit of analysis was the first ICU stay of each hospital admission (the lowest unit-visit number within each patienthealthsystemstayid). The six acute brain-injury phenotypes were assigned from the eICU diagnosis table using the identical ICD-9 and ICD-10 prefix lists and priority hierarchy of the primary analysis (eTable 1), applied to both the ICD-9 and ICD-10 codes recorded in the icd9code field. Command-following was the Glasgow Coma Scale motor subscore of 6 ("Obeys") charted in nurseCharting (cell label "Glasgow coma score", value name "Motor"); stays with fewer than three motor examinations were excluded; and a baseline non-follower had a first motor examination below 6 within 12 hours of unit admission. Time was measured from unit admission. The competing events were in-unit death (unitdischargestatus = ”Expired”) and alive unit discharge without recovery, with observation otherwise censored at the unit-stay end. The Aalen-Johansen estimator, the 400-resample bootstrap, and the instability metrics were the same in-house implementations used for the primary cohort. Of 12,397 brain-injury first stays, 7,615 (61.4%) had three or more motor examinations; this lower retention than in MIMIC-IV (97.4%) reflects that the motor subscore is charted less consistently than the total Glasgow Coma Scale in eICU. Results are in eTable 17.

#### S2. Supplementary Tables

**eTable 1. Diagnosis code lists for the six acute brain-injury phenotypes.** Codes are matched on the recorded prefix within the stated ICD version.

| Phenotype | ICD-9 prefix | ICD-10 prefix |
|---|---|---|
| Subarachnoid hemorrhage | 430 | I60 |
| Intracerebral hemorrhage | 431 | I61 |
| Subdural hemorrhage | 432 | I62 |
| Acute ischemic stroke | 433, 434 | I63 |
| Traumatic brain injury | 800 to 804, 850 to 854 | S06 |
| Anoxic or hypoxic-ischemic | 348.1, 348.5 | G93.1, G93.82 |

**eTable 2. Glasgow Coma Scale motor value set and numeric mapping.** Command-following corresponds to the highest value. Values were taken from nursing flowsheet documentation (item 223901).

| Charted motor value | Numeric score | Classification |
|---|---|---|
| Obeys Commands | 6 | Command-following |
| Localizes Pain | 5 | Non-following |
| Flex-withdraws | 4 | Non-following |
| Abnormal Flexion | 3 | Non-following |
| Abnormal extension | 2 | Non-following |
| No response | 1 | Non-following |

**eTable 3. Cumulative incidence of command-following by phenotype among baseline non-followers.** Values are the three-state Aalen-Johansen cumulative incidence (%) of a first command-following examination, with the competing cumulative incidence of death or hospice at 14 days shown in the final column. These are from the same three-state model as eTable 12 and Figure 2; baseline non-followers per phenotype are shown as n.

| Phenotype | n | 24 h | 72 h | 7 d | 14 d | Death or hospice, 14 d |
|---|---|---|---|---|---|---|
| Acute ischemic stroke | 1476 | 53.1 | 66.3 | 73.9 | 77.2 | 17.2 |
| Traumatic brain injury | 1419 | 54.8 | 64.5 | 69.8 | 73.8 | 19.0 |
| Subdural hemorrhage | 237 | 43.5 | 59.1 | 63.7 | 65.8 | 25.7 |
| Intracerebral hemorrhage | 1094 | 38.2 | 50.4 | 55.6 | 59.3 | 34.6 |
| Subarachnoid hemorrhage | 533 | 35.6 | 49.3 | 53.7 | 58.3 | 37.1 |
| Anoxic or hypoxic-ischemic | 739 | 16.4 | 25.1 | 33.5 | 36.9 | 52.3 |

**eTable 4. Instability of recovery by phenotype.** Among patients who reached command-following at least once. Transient denotes a first recovery lost at the next examination; in-and-out denotes at least one crossing back below the threshold after recovery; crossings is the per-patient

median number of threshold crossings.

| Phenotype | n | Transient, % | Sustained, % | In-and-out, % | Crossings, median |
|---|---|---|---|---|---|
| Acute ischemic stroke | 1139 | 21.2 | 63.7 | 53.2 | 2.0 |
| Traumatic brain injury | 1046 | 21.2 | 55.8 | 61.4 | 3.0 |
| Intracerebral hemorrhage | 649 | 24.5 | 51.6 | 74.4 | 4.0 |
| Subarachnoid hemorrhage | 311 | 25.1 | 64.6 | 79.7 | 5.0 |
| Subdural hemorrhage | 156 | 18.6 | 59.0 | 59.0 | 2.5 |
| Anoxic or hypoxic-ischemic | 272 | 23.5 | 54.8 | 58.8 | 2.0 |

The proportion with a transient first recovery did not differ across phenotypes (Cramer V, 0.04; adjusted $P = .32$). The number of crossings differed across phenotypes (epsilon-squared, 0.05; adjusted $P < .001$). Each additional crossing was associated with lower odds of discharge home (odds ratio, 0.83; 95% CI [0.80, 0.85]; adjusted $P < .001$).

**eTable 5. Sedation share of non-following examinations and the sedation contrast, by phenotype.** Sedation share is the percentage of non-following examinations recorded during an active continuous sedative or analgesic infusion.

| Phenotype | Non-following exams during sedation, % |
|---|---|
| Subdural hemorrhage | 32.2 |
| Intracerebral hemorrhage | 33.8 |
| Acute ischemic stroke | 35.9 |
| Subarachnoid hemorrhage | 40.6 |
| Traumatic brain injury | 44.5 |
| Anoxic or hypoxic-ischemic | 44.8 |

Across the cohort, 39.1% of non-following examinations coincided with active sedation. An examination during active sedation was more likely to show non-following than one outside sedation (relative risk, 2.44; 95% CI [2.35, 2.53]; risk difference, +0.42; 95% CI [+0.41, +0.44]). In the 1,702 baseline non-followers who never received any continuous sedative or analgesic infusion, the cumulative incidence of command-following at 72 hours was 47.5%, lower than the 55.1% in the full cohort (eFigure 2).

**eTable 6. Measurement-probe model card.** The probe predicts sustained command-following before discharge among baseline non-followers who survived at least 24 hours. It is a measurement description, not a deployable clinical tool.

| Field | Value |
|---|---|
| Task | Binary classification of sustained command-following before discharge |
| Model | Gradient-boosted decision trees (HistGradientBoostingClassifier, scikit-learn 1.6) |
| Sample | 5,027 baseline non-followers surviving at least 24 hours; outcome prevalence 41.4% |
| Features (first 24 h only) | First, last, minimum, maximum, and slope of the motor score; number of examinations; any command-following in the window; fraction of examinations during sedation; any neuromuscular blockade; mean Richmond Agitation-Sedation Scale value; age; phenotype |
| Baseline comparator | Logistic regression on age and phenotype only |
| Validation | 5-fold stratified cross-validation, out-of-fold predictions |
| Discrimination | Area under the curve 0.82 (95% CI [0.81, 0.83]); baseline 0.64 (95% CI [0.63, 0.66]) |
| Difference vs baseline | +0.18 (95% CI [+0.16, +0.19]); paired bootstrap $P < .001$ |
| Calibration | Brier score 0.17 (model), 0.23 (baseline) |

| Field | Value |
|---|---|
| External validation | None |

**eTable 7. Adjusted hazards of command-following and of death.** Cause-specific hazard ratios (csHR; Cox model, competing events censored) and subdistribution hazard ratios (SHR; Fine-Gray model) for the command-following event, and cause-specific hazard ratios for the competing death event, among baseline non-followers (ischemic stroke is the reference phenotype). Both recovery models adjust for the listed covariates simultaneously.

| Covariate | Recovery csHR [95% CI] | Recovery SHR [95% CI] | Death csHR [95% CI] |
|---|---|---|---|
| Traumatic brain injury | 1.02 [0.93, 1.11] | 1.05 [0.97, 1.13] | 1.35 [1.13, 1.60] |
| Intracerebral hemorrhage | 0.72 [0.66, 0.80] | 0.75 [0.69, 0.82] | 1.65 [1.41, 1.93] |
| Subarachnoid hemorrhage | 0.67 [0.59, 0.76] | 0.72 [0.64, 0.81] | 1.71 [1.43, 2.06] |
| Subdural hemorrhage | 0.94 [0.79, 1.11] | 0.90 [0.77, 1.05] | 1.88 [1.42, 2.48] |
| Anoxic or hypoxic-ischemic | 0.31 [0.27, 0.35] | 0.41 [0.37, 0.46] | 1.60 [1.37, 1.88] |
| Age, per 10 years | 0.98 [0.96, 1.00] | 0.96 [0.95, 0.98] | 1.16 [1.12, 1.19] |
| Female sex | 0.94 [0.88, 1.01] | 0.93 [0.88, 1.00] | 1.08 [0.98, 1.20] |
| Sedated at baseline | 1.43 [1.32, 1.55] | 1.46 [1.36, 1.57] | 0.65 [0.58, 0.73] |
| Mechanically ventilated | 0.73 [0.67, 0.80] | 0.83 [0.77, 0.90] | 0.98 [0.86, 1.11] |

The subdistribution hazard ratios are from a Fine-Gray model in which both death or hospice and alive ICU discharge without recovery are treated as competing events (consistent with the descriptive three-state cumulative incidence functions); only administrative censoring at 14 days enters the inverse-probability-of-censoring weights. Death cause-specific hazard ratios are shown to display the competing process; the recovery and death hazards move in opposite directions across phenotypes, which is why the unadjusted cumulative incidence is best read through the competing-risks estimator rather than a single survival curve.

**eTable 8. Predictive-probe performance for sustained command-following.** Discrimination, calibration, and an operating point for the three models among baseline non-followers surviving at least 24 hours. The local language model was evaluated zero-shot on a held-out random sample; the same-sample column reports each model restricted to those patients for a fair comparison.

| Model | n | AUROC [95% CI] | Brier | Calibration slope | Sensitivity | Specificity | AUROC, same 700-patient sample |
|---|---|---|---|---|---|---|---|
| Gradient boosting (tabular) | 5,027 | 0.82 [0.81, 0.83] | 0.17 | 0.92 | 0.83 | 0.68 | 0.79 |
| Deep temporal model (GRU) | 5,027 | 0.79 [0.78, 0.80] | 0.18 | 0.98 | 0.72 | 0.74 | 0.77 |
| Local LLM (Qwen3.5-4B, zero-shot) | 700 | 0.75 [0.71, 0.78] | 0.23 | 0.19 | 0.74 | 0.70 | 0.75 |

The gradient-boosted model over tabular first-day summaries gave the best discrimination; the deep temporal model over the raw examination sequence did not improve on it, indicating that the predictive signal is largely captured by simple summaries of the first day. The zero-shot language model, reading only the flowsheet text with no fine-tuning, reached an area under the curve of 0.75 on the same patients, approaching the trained models but with poor calibration (calibration slope 0.19), as expected for an uncalibrated zero-shot reader. In the next-examination forecasting task the recurrent network reached an area under the curve of 0.965 against a first-order Markov baseline of 0.929, with next-state persistence probabilities of 0.94 (following to following) and 0.08 (not following to following), indicating that the bedside fluctuation is highly structured over the short term and accumulates into multiple threshold crossings only across the longer stay.

**eTable 9. Non-following examinations by sedation class, depth, ventilation, and time since discontinuation.** Among baseline non-followers. For each contrast the percentage of examinations that were non-following is shown for the exposed and reference groups, with the number of examinations in each group and the relative risk from a stay-clustered bootstrap. Reference groups are stated explicitly; sedative and opioid infusions can overlap, so each drug-class contrast is defined against the absence of that class.

| Contrast (reference) | Exposed % (n exams) | Reference % (n exams) | Relative risk [95% CI] |
|---|---|---|---|
| Deep sedation, RASS -4/-5 (vs RASS > -4) | 98.0 (80,860) | 28.1 (451,046) | 3.49 [3.37, 3.61] |
| Active invasive ventilation (vs none) | 74.0 (238,833) | 19.8 (456,520) | 3.73 [3.55, 3.92] |
| Active hypnotic infusion (vs no hypnotic) | 71.9 (133,634) | 30.4 (561,719) | 2.36 [2.29, 2.45] |
| Active opioid infusion (vs no opioid) | 76.7 (54,336) | 35.2 (641,017) | 2.18 [2.11, 2.26] |
| Within 6 h of stopping a hypnotic (vs >6 h off, off-hypnotic only) | 51.8 (38,569) | 41.0 (204,461) | 1.26 [1.22, 1.31] |

The non-following share rose monotonically with sedation depth, from 6.8% at RASS 0 to 85.2% at RASS -3, 97.1% at RASS -4, and 99.3% at RASS -5 (Figure 4a). Sedation-practice subgroups recovered in opposite directions (cumulative incidence of command-following at 72 hours, three-state model): full cohort 55.1%; never deeply sedated (n = 2,065) about 75%; never given any continuous sedative or analgesic (n = 1,702) about 47%. The opposite direction of these two subgroups indicates that sedation practice marks different populations; the contrasts are descriptive, not causal.

**eTable 10. Neuromuscular blockade and the primary handling.** Examinations during active blockade were excluded in the primary analysis.

| | Value |
|---|---|
| Examinations during blockade | 6,020 of 695,353 (0.87%) |
| Patients with any blockade examination | 454 |
| Share of examinations by phenotype | anoxic 3.3%, SAH 1.0%, TBI 0.8%, AIS 0.7%, ICH 0.4%, SDH 0.2% |
| Baseline non-followers (blockade excluded, sensitivity) | 5,483 (vs 5,498 carried forward, primary) |
| Command-following at 72 h | 55.8% (blockade excluded) vs 55.1% (carried forward, displayed) |
| Transient first recovery | 22.1% (blockade excluded) vs 22.2% (carried forward) |

**eTable 11. Threshold crossings adjusted for examination frequency.** Negative-binomial incidence-rate ratios for the number of crossings with an offset for the number of examinations (per-examination rate) and, separately, for ICU days (per-day rate), relative to ischemic stroke. The raw-count incidence-rate ratios (no offset) are shown for contrast.

| Phenotype | Raw IRR [95% CI] | Per-examination IRR [95% CI] | Per-ICU-day IRR [95% CI] |
|---|---|---|---|
| Traumatic brain injury | 1.21 [1.08, 1.36] | 0.96 [0.87, 1.06] | 1.41 [1.28, 1.57] |
| Intracerebral hemorrhage | 1.63 [1.47, 1.82] | 0.78 [0.70, 0.88] | 1.23 [1.10, 1.38] |
| Subarachnoid hemorrhage | 2.49 [2.17, 2.85] | 0.65 [0.56, 0.74] | 1.07 [0.93, 1.23] |
| Subdural hemorrhage | 1.23 [1.00, 1.50] | 0.82 [0.68, 1.00] | 1.22 [1.00, 1.48] |
| Anoxic-ischemic | 1.09 [0.93, 1.27] | 0.98 [0.84, 1.15] | 0.94 [0.80, 1.10] |

The raw count is higher in the hemorrhagic phenotypes, but this reflects more frequent examination; per examination, subarachnoid and intracerebral hemorrhage have fewer crossings than ischemic stroke. The median crossing rate per 10 examinations was similar across phenotypes (ischemic stroke 0.86, traumatic 0.73, anoxic 0.71, subdural 0.61, intracerebral 0.59, subarachnoid 0.51).

**eTable 12. Three-state competing-risks cumulative incidence.** Modeling alive ICU discharge without recovery as a separate competing event, among baseline non-followers.

| State | 24 h | 72 h | 7 d | 14 d |
|---|---|---|---|---|
| Command-following | 43.5 | 55.1 | 61.4 | 65.0 |
| Death or hospice | 4.6 | 15.3 | 23.2 | 28.2 |
| Alive discharge without recovery | 0.8 | 2.9 | 4.3 | 6.8 |
| (sum) | 48.9 | 73.3 | 88.9 | 100.0 |

**eTable 13. Phenotype co-occurrence and single-phenotype sensitivity.** A single primary phenotype was assigned by a fixed hierarchy when more than one brain-injury code was present.

| | Value |
|---|---|
| Hospitalizations with a brain-injury code | 29,292 |
| Hospitalizations with two or more phenotypes | 3,344 (11.4%) |
| Single-phenotype baseline non-followers | 4,147 |
| Command-following at 72 h, single-phenotype only | 57.6% |
| Command-following at 72 h, all | 55.1% |

**eTable 14. Exploratory association between threshold crossings and discharge home.** Logistic regression odds ratio per additional crossing, with progressively fuller adjustment.

| Model | OR per crossing [95% CI] |
|---|---|
| Adjusted for age and phenotype | 0.83 [0.80, 0.85] |
| Fuller adjustment (age, phenotype, vasopressor, sedation exposure, length of stay) | 0.90 [0.87, 0.94] |

| | |
|---|---|
| Fuller adjustment plus mechanical ventilation | 0.90 [0.87, 0.94] |

The association attenuates with fuller adjustment but persists. Because discharge disposition is influenced by social and systemic factors not captured here, this analysis is exploratory and is not a prognostic claim.

**eTable 15. Sensitivity of the command-following endpoint.** Cumulative incidence (%) among baseline non-followers. First command-following is contrasted with sustained command-following (onset of the final uninterrupted following period held to discharge) and a two-consecutive-examination definition; the interpretable-only row re-derives the endpoint using examinations free of neuromuscular blockade, deep sedation, and active hypnotic infusion; the competing event is split into in-hospital death and discharge to hospice or comfort care.

| Endpoint | 24 h | 72 h | 7 d | 14 d |
|---|---|---|---|---|
| First command-following | 43.7 | 55.3 | 61.5 | 65.2 |
| Sustained command-following (held to discharge) | 23.8 | 33.8 | 42.1 | 52.1 |
| Two consecutive following examinations | 39.9 | 51.2 | 57.9 | 61.7 |
| First command-following, interpretable examinations only (n = 3,518) | 32.2 | 48.3 | 58.4 | 65.3 |
| Competing event: in-hospital death (n = 1,867) | 4.3 | 14.1 | 21.5 | 26.1 |
| Competing event: hospice or comfort-care discharge (n = 212) | 0.3 | 1.1 | 1.5 | 1.9 |

The median time to sustained command-following was 32 hours, against 12.2 hours to first command-following. The competing event was almost entirely in-hospital death rather than an early comfort-care transition, and the interpretable-only curve was lower early (the early recovery is partly pharmacological) but reached the same 14-day value.

**eTable 16. Recovery of command-following by baseline Glasgow Coma Scale motor**

**category.** Cumulative incidence (%) of first command-following among baseline non-followers, by the worst (first) motor response within 12 hours of admission.

| Baseline motor response | n | 24 h | 72 h | 7 d | 14 d |
|---|---|---|---|---|---|
| Localizes pain | 1784 | 57.7 | 71.3 | 76.6 | 79.6 |
| Flexion-withdrawal | 1165 | 40.4 | 54.8 | 63.0 | 66.5 |
| No response | 2124 | 38.7 | 48.3 | 54.5 | 58.6 |
| Abnormal flexion | 255 | 19.7 | 24.8 | 29.5 | 34.6 |
| Abnormal extension | 154 | 12.3 | 20.1 | 25.3 | 29.9 |

Patients who localized to pain recovered most often and posturing (abnormal flexion or extension) least often; patients with no motor response recovered more often than those with flexor or extensor posturing.

**eTable 17. External validation in the eICU Collaborative Research Database (208 U.S. hospitals).** The command-following construct (Glasgow Coma Scale motor score of 6), the six ICD-based phenotypes, the three-or-more-examination requirement, the 12-hour baseline-non-follower rule, the Aalen-Johansen estimator, and the instability metrics were held identical to the primary analysis (eMethods S1.9). MIMIC-IV values are reproduced from the main analysis for comparison.

*Panel A. Cumulative incidence (%) of first command-following among baseline non-followers.*

| Cohort | Hospitals | Non-followers, n | 24 h | 72 h | 7 d | 14 d | Median time, h |
|---|---|---|---|---|---|---|---|
| MIMIC-IV | 1 | 5498 | 43.5 | 55.1 | 61.4 | 65.0 | 12.2 |
| eICU-CRD | 208 | 2519 | 35.1 | 51.3 | 60.0 | 64.8 | 16.2 |

*Panel B. Fourteen-day cumulative incidence (%) of command-following by phenotype.*

| Phenotype | MIMIC-IV | eICU-CRD (n) |
|---|---|---|
| Acute ischemic stroke | 77.2 | 79.3 (567) |
| Traumatic brain injury | 73.8 | 70.3 (840) |
| Subdural hemorrhage | 65.8 | 65.1 (583) |
| Intracerebral hemorrhage | 59.3 | 54.1 (99) |
| Subarachnoid hemorrhage | 58.3 | 53.3 (161) |
| Anoxic or hypoxic-ischemic | 36.9 | 29.1 (269) |

*Panel C. Instability of recovery among patients who reached command-following at least once.*

| Metric | MIMIC-IV | eICU-CRD |
|---|---|---|
| Patients who recovered, n | 3573 | 1479 |
| Transient first recovery, % | 22.2 | 22.7 |
| Crossed threshold at least once after recovery, % | 62.4 | 64.2 |
| Threshold crossings, median | 3 | 3 |

The 14-day cumulative incidence of command-following (64.8% versus 65.0%), the rank order of phenotypes (lowest in anoxic injury, highest in ischemic stroke), and the instability of recovery (transient first recovery 22.7% versus 22.2%; median 3 crossings) reproduced in the multicenter cohort. Early incidence was lower in eICU-CRD, consistent with sparser first-day motor documentation and shorter unit stays; the competing 14-day cumulative incidence of in-unit death among non-followers was 26.0%. Phenotype cells with fewer than 200 patients (intracerebral hemorrhage, subarachnoid hemorrhage) are reported for completeness but are less precise.

#### S3. Supplementary Figures

**eFigure 1. Instability of command-following after it first appears.** (a) Among examinations performed after a patient's first command-following, the percentage still showing command-following, by time since first recovery (slate line with bootstrap 95% confidence band); the dashed line marks the expectation if recovery were a stable acquired state. Command-following was confirmed at only 78% of immediately following examinations and was present at about 60% of examinations over the following week. (b) All 3,573 patients who recovered command-following, one per row, sorted by time to first command-following, with dark slate denoting following commands, pale gray not following, and white after discharge or death; the smooth boundary is the timing of first recovery and the gray within the slate region is relapse. (Rendered as efig_trajectories.pdf.)

**eFigure 2. Predictive-probe performance.** Receiver operating characteristic curves, calibration, decision-curve net benefit, and next-examination forecasting for the gradient-boosted, deep temporal, and zero-shot language models. (Rendered as fig6_models.pdf.)

#### S4. Code and Data Availability

MIMIC-IV and the eICU Collaborative Research Database are both available to credentialed users through PhysioNet under a data use agreement; no individual-level data are included in this Supplement or the associated repository. The analysis code, including the cohort build, the in-house Aalen-Johansen estimator, the figure builders, and the eICU external-validation pipeline (60_eicu_extract.py, 61_eicu_validation.py), is available at https://github.com/Alon-Gorenshtein/study_command_following. The single source of truth for every value reported here is the set of analysis digests emitted by the pipeline: the three-state competing-risks digest (threestate_digest.json) for the cumulative incidence functions, stats_digest.json and the advancedstatistics digest for the adjusted hazard and rate models, and results_digest.json for the cohort summaries.
